# Heterogeneity impacts biomarker discovery for precision medicine

**DOI:** 10.1101/2022.02.14.22270972

**Authors:** Kenneth Smith, Sharlee Climer

## Abstract

Precision medicine is advancing patient care for complex human diseases. Discovery of biomarkers to diagnose specific subtypes within a heterogeneous diseased population is a key step towards realizing the benefits of precision medicine. However, popular statistical methods for evaluating candidate biomarkers – fold change (FC) and area under the receiver operating characteristic curve (AUC) – were designed for homogeneous data. Herein, we evaluate the performance of these metrics in heterogeneous populations. Using simulated biomarkers that are nearly ‘ideal’ for distinguishing subgroups of various proportions of the diseased population, we observe that AUC misses all up to subset size of 50% and FC misses all biomarkers entirely. We introduce a simple new measure to address this shortfall and run a series of trials comprised of simulated and biological data to demonstrate its utility for evaluating biomarkers associated with disease subtypes.

## Introduction

Advances in precision medicine (PM) for cancer patients is extending the healthspan for countless lives by tailoring treatments to heterogeneous cancer subtypes. PM utilizes specific biomarker information, such as genetics and levels of proteins being produced, to diagnose their specific subtype of the disease and enable tailored treatments, prognoses, and monitoring. An additional benefit of PM is that it facilitates understanding of underlying biological mechanisms by teasing apart biomarkers into subtype groups. Knowledge of distinct biomarkers associated with each subtype empowers drug discovery as well as selections of individuals for drug trials. Many complex diseases are heterogeneous and hold potential to benefit from PM. For example, heterogeneous subtypes of late-onset Alzheimer disease (AD) are exhibited by the spectrum of genetic and environmental risk factors and clinical outcomes observed for this enigmatic disease. Efforts are underway to enable PM for AD, including the Accelerating Medicines Partnership® for AD 2.0^1^, which began in 2021, and Alzheimer Precision Medicine Initiative^2^, which began in 2016.

The realization of successful PM can only be attained by identifying disease subtypes and developing practical methods to diagnose and treat each subtype. A common approach is to use statistical methods to test associations of candidate biomarkers with the disease. Candidate biomarkers may include genetics, demographics, lifestyle, and a host of observations such as imaging data, body mass index, and omics data – which may include levels of gene expression, proteins, lipids, and metabolites. Different statistics are used for categorical, ordinal, and numerical data types. Herein we focus on numerical data types, which includes omics data, measurements from imaging data (such as PET amyloid load), and other observations that are quantified as numerical values. Popular statistics for this domain include fold change (FC) of levels of candidate biomarkers between diseased cases and normal controls and area under the receiver operating characteristic curve (AUC)^3^. In addition to tests on individual biomarkers, patterns comprised of multiple biomarkers can be identified by modeling the data as a network in which biomarkers are represented by nodes and correlations between pairs of biomarkers are represented by edges between the corresponding node pairs^4–9^. Clusters of intercorrelated biomarkers are identified and evaluated using methods such as modularity^7,10^ or gene enrichment^11^.

This nascent research field faces challenges due to multiple issues, such as the need for large sample sizes for studies to elicit power to sift out a subtype that may only represent a small fraction of the diseased cases. Another major challenge is that traditional statistical methods that are successful for global biomarkers can be inappropriate for subset biomarker identification, and each approach needs to be reevaluated for use in this distinct domain. We previously reported that the use of standard correlation metrics in network modeling leads to increased type II errors in the presence of subtype groups^8,9,12,13^. All the correlation measures that we have examined, including Pearson’s correlation coefficient^14^, *r*-squared^15^, dot product^16^, and mutual information^17^, return single scalar values that are crippled by heterogeneity^9^. They are universal measures, in that individuals in the entire group are viewed as a whole, and thus subtle but crucial subgroup structures are obscured. If two analytes are highly correlated for a subset of individuals but not at all correlated for the others, the correlation value is reduced due to the latter individuals, thereby contributing to false negative signals^8,9^.

In this manuscript, we examine the use of FC and AUC when subtype groups exist. A traditional approach for identifying differentially expressed analytes involves calculating the FC of the analyte expression levels between two groups as a quotient: (level in diseased cases) / (level in normal controls). If the quotient is above or below a given cutoff, the analyte is considered differentially expressed. A single value representing the expression level of the analyte is required for each group; usually the median or mean. Typically, a cutoff of >2 is used to indicate significant up-regulation in the diseased cases group and a cutoff of <0.5 for down-regulation. In order to more easily interpret across both up- and down-regulated analytes, the log2FC is often employed, where log2FC = abs(log_2_((level in diseased cases) / (level in normal controls))), providing a cutoff of log2FC > 1 tests for both up- and down-regulation^18^. FC calculations are unstable when the expression levels are near the noise level of the measurement system. This can lead to false positives at low intensity levels. At the other end of the spectrum, FC is also biased against samples that have high expression levels, but small differences between two groups^19^. Mariani *et. al*. reported that high FC cutoffs are needed for low intensity genes and lower cutoffs are needed for high intensity genes. They introduced a variable FC cutoff-based approach that uses LOESS to estimate a variance based on expression intensity, thereby alleviating the bias at both high and low intensity levels^19^. Despite these improvements to the FC calculation, there is a fundamental problem with this metric: Use of the mean or median in the presence of heterogeneity tends to miss subgroup signals, as demonstrated in this manuscript.

Standard 2 × 2 contingency tables are commonly used to assess predictive accuracy of biomarkers using various statistics, such as sensitivity/specificity, precision/recall, Fisher’s Exact Test^20^, and Youden’s J index^21^. Note that Youden’s J definition can be rearranged to produce a simple interpretation: J = TPR – FPR, where TPR is the true positive rate and FPR is the false positive rate. A key benefit of utilizing Youden’s J is that subgroups are captured, rather than being lost in a summary statistic, as is done with FC. However, without other information, subgroups may be overlooked due to the existence of moderate case/control biomarkers with the same J value, just higher TPR and FPR, e.g. J = 0.20 - 0.01 vs J = 0.70 - 0.51.

When testing numerical values, 2 × 2 contingency tables require the selection of a threshold to separate diagnostic classifications. A key strength of AUC is that it has no reliance upon a specified threshold to predict diseased cases vs. normal controls. This metric originated as a tool for radar receivers, spread throughout engineering and medical domains, and has become a prevalent tool for evaluating the diagnostic ability of biomarkers^3,22,23^. AUC simultaneously accounts for sensitivity and specificity across all threshold values as a plot of the TPR vs. FPR is constructed and the area under the curve is returned as the AUC value^24^. The plot for a random classifier would tend toward a diagonal line from (0,0) to (1,1) with an AUC value of 0.5. A ‘perfect’ predictor would have FPR=0 and TPR=1 for all thresholds of the biomarker and a corresponding AUC value of 1. An example of this rare event was reported by Karikari et al. for discriminating Alzheimer’s disease from healthy young adults using plasma tau phosphorylated at threonine 181 (pTau-181)^25^.

There isn’t a consensus for a significance cutoff for AUC values. Previous publications have suggested an AUC between 0.7 and 0.8 as acceptable and greater than 0.8 as excellent^26,27^, while the National Center on Response to Intervention’s Technical Standard sets AUC values between 0.75 and 0.85 as ‘partially convincing’ and below 0.75 as ‘unconvincing’^28^. On the other hand, it has been recommended that no set value should be utilized; rather AUC values should be used to compare predictors within a single domain rather than enforcing a strict cutoff value^29–32^.

In addition to evaluating biomarkers across all threshold values, AUC has several other beneficial properties. It is a simple and intuitive measure and the corresponding ROC plot provides additional information beyond the scalar value. Also, there are no parameters to be tuned, yielding robust reproducibility.

There are also some well-known issues with AUC. First, small sample size can yield poor performance^33,34^. Second, AUC includes the areas under the ROC curve that represent threshold values that are not utilized in practical applications^35^. A related issue is when the ROC curves of two different biomarkers cross, the relative AUC values may be misleading^36^.

In general, the points in the ROC curve arise *solely* from differences in TPR and FPR and are not scaled across threshold values, resulting with the possibility of incremental spans of threshold values being stretched across broad regions of the area under the curve. In clinical practice, target thresholds or threshold ranges are used to flag individuals at risk. AUC values are generally computed over clean data that have been acquired and processed using highly uniform methods, but this uniformity deteriorates when moving from bench to bedside. Consequently, examination of AUC values and plots do not directly provide insights for evaluating biomarkers for robustness across measurement error. The metric introduced in this manuscript addresses this issue.

The AUC metric is entirely dependent upon, and equally weighted on, the TPR and FPR. When testing across a heterogeneous group, the TPR for a perfect biomarker has an upper limit equal to the proportion of the subtype. In general, the percentage of samples representing subtype *i, p*_*i*_, produces a corresponding upper bound on the TPR and lower bound of 1 - *p*_*i*_ on the false negative rate. Consequently, we hypothesized that screening based on AUC may discard valuable subtype biomarkers, regardless of sample size. Using simulated tests mimicking nearly ‘ideal’ biomarkers for subsets of disease cases, we demonstrate the failure of AUC to capture their significance.

In this manuscript, we introduce a metric for evaluating biomarkers of heterogeneous phenotypes which springs from observations of the *distributions* of values, rather than TPR, FPR, and other traditional metrics. Consider a biomarker that is a strong indicator of a subset of diseased cases, referred to as associated cases. We assume here that the cases that are not associated exhibit biomarker levels that are similar to the normal controls. Consequently, the distribution of biomarker levels for the cases tends to skew or exhibit a bimodal profile, where one of the modes lines up with the controls’ distribution. It should be noted that normal controls might show a bimodal distribution also. For example, blood sugar levels are high following a meal and low just before a meal, so controls sampled at varying times of day would be prone to exhibit a bimodal curve for this analyte.

Aiming to identify aberrant bimodal distributions, we propose a metric which calculates the difference between the bimodalities of the diseased cases and normal controls. A number of formulae for distinguishing between unimodality and bimodality have been proposed and evaluated^37^. Hartigan’s Dip Statistic (HDS)^38^ and the Bimodality Coefficient (BC)^39^ have both been shown to have good accuracy to detect bimodality^37^. Note that high skewness in a unimodal distribution tends to increase BC values and can lead to false-positive bimodal predictions^40^. We selected BC as we are interested in identifying either bimodality or skewness that is significantly different between cases and controls.

BC was introduced by SAS in 1990 and is based on three parameters of the array of values: cardinality (*n*), skewness (*s*), and kurtosis (*k*). The value is computed as follows:

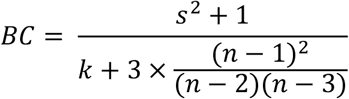

BC values range from zero to one and a uniform distribution has a value of 5/9 ≈ 0.555. Higher values indicate greater bimodality.

We propose the following measure, bimodality coefficient difference (BCD), for identifying biomarkers representing subtypes in heterogeneous populations:

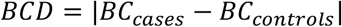

The absolute value is applied as a protective factor may result with the controls having a higher BC value than the cases. BCD can range from zero to one, but we observe from our trials that relatively low values indicate significance. Using a series of simulation trials and biological data, we demonstrate the effectiveness of this metric for identifying promising biomarkers representing subsets of cases.

## Methods

### Biological data trials

We utilized publicly-available gene expression data from human cortex tissue generated using Sentrix HumanRef-8 Expression BeadChip^41^. These data are available on NCBI’s Gene Expression Omnibus (GEO), Accession GSE15222. Standard protocols for cRNA hybridization and BeadStudio software, with Illumina’s custom error model, were utilized in data generation, as previously described^41^. Data for 8,650 genes for 176 AD cases and 187 controls are provided and used for the current study.

### Simulated data trials

In our simulations, samples were drawn from one of two normal distributions, N_1_ and N_2_, with the following means and standard deviations: N_1_∼(0.03, 0.04) and N_2_∼(0.40, 0.016). These means and standard deviations were derived from analysis of highly differentially-expressed proteins from our COVID-19 study (**Supplementary Information**). The size of the subtype, as a percentage of the cases, was varied over seven scenarios from 0% to 50%. In each scenario, the cases in the subtype group were sampled from N_2_ and the remaining cases, along with all controls, were sampled from N_1_. A total of 1000 cases and 1000 controls were simulated in each trial. Each scenario was tested using 1000 trials. Histograms for randomly selected trials are shown in **Figure 1**.

**Figure 1.**
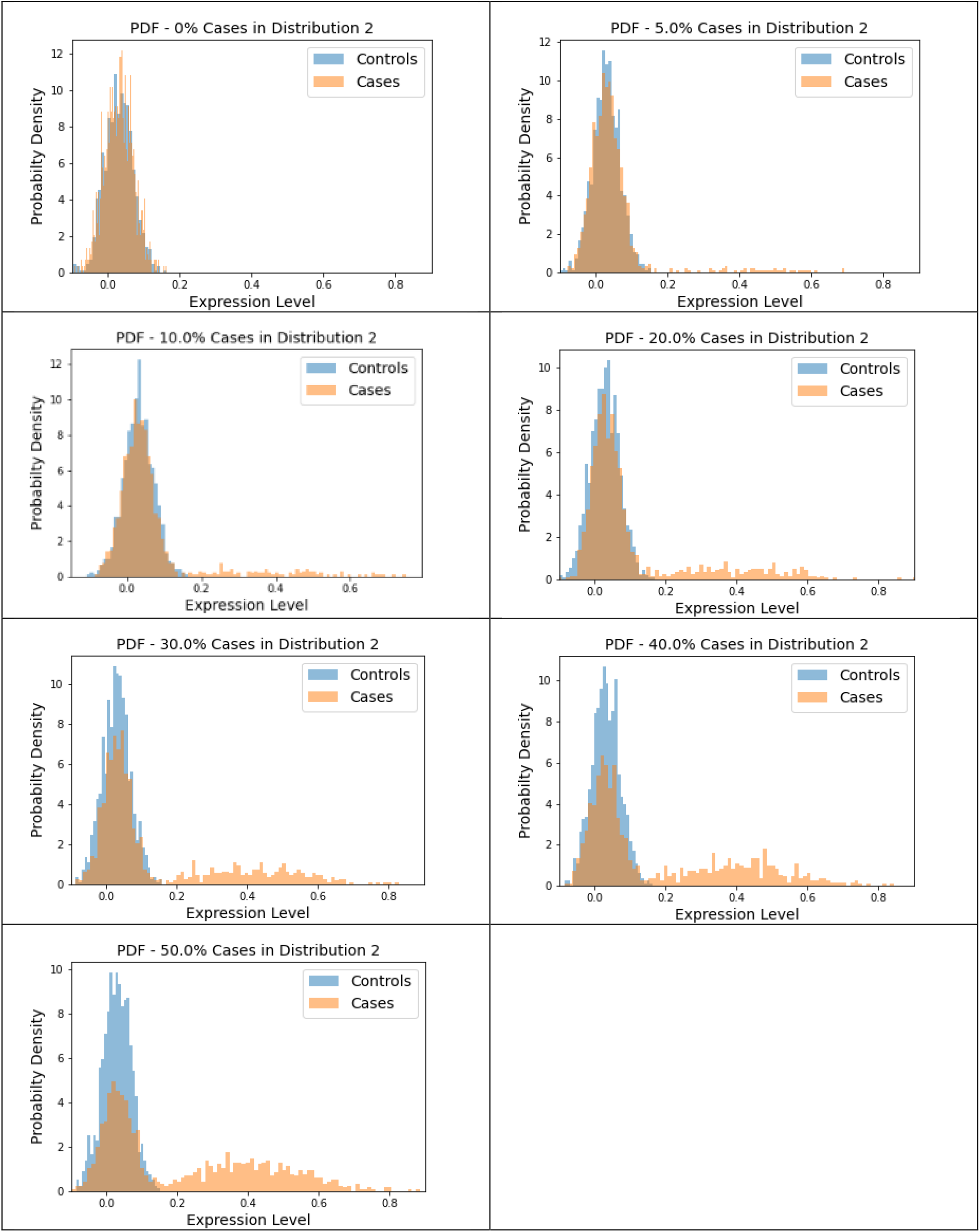
Sample histograms for the simulation trials. Shown are random histograms drawn from the 1000 trials for each of the seven subset size scenarios.

### Data pre-processing

The AD data were pre-processed by the Myers’ lab, as described previously^41^. We utilized these data without further normalizations in order to preserve the natural distributions. However, outliers disproportionately affect BC values, so they were trimmed by replacing values more/less than 3 interquartile ranges above/below the third/first quartile with the value that is equal to 3 interquartile ranges above/below the third/first quartile.

## Results

### Simulated Trials

We generated large-scale simulated data for a total of 7000 pseudo analytes and analyzed each using FC, AUC, and BCD. The subset size of zero provides a baseline for which no association should be observed as all of the data points for cases and controls are drawn from the N_1_ distribution. Results for the simulations are summarized in **Table 1**. Sample ROC curves for each scenario are shown in **Figure 2**.

**Table 1.** Median values for the simulation trials, with minimum and maximum values shown in brackets. The first row represents no subsets, where all of the cases and controls values are drawn from the N_1_ distribution. Subsequent rows represent trials with 5%, 10%, 20%, 30%, 40%, and 50%, respectively, of the cases values drawn from the N_2_ distribution and represent the disease subtype. For each scenario, 1000 cases and 1000 controls values were generated for each of 1000 trials.

| **Trial** | **log2FC** | **AUC** | **BCD** |
| --- | --- | --- | --- |
| *Sim_0%* | 0.016 [5.47E-06, 0.080] | 0.508 [0.491, 0.542] | 0.016 [8.68E-06, 0.076] |
| *Sim_5%* | 0.0276 [2.25E-06, 0.107] | 0.525 [0.483, 0.563] | 0.145 [0.066, 0.214] |
| *Sim_10%* | 0.0544 [2.79E-04, 0.143] | 0.548 [0.509, 0.605] | 0.282 [0.214, 0.350] |
| *Sim_20%* | 0.124 [0.037, 0.210] | 0.598 [0.563, 0.630] | 0.395 [0.325, 0.458] |
| *Sim_30%* | 0.206 [0.071, 0.320] | 0.646 [0.617, 0.675] | 0.441 [0.372, 0.514] |
| *Sim_40%* | 0.337 [0.134, 0.452] | 0.695 [0.669, 0.723] | 0.464 [0.389, 0.525] |
| *Sim_50%* | 0.608 [0.221, 0.821] | 0.743 [0.719, 0.768] | 0.438 [0.383, 0.504] |

**Figure 2.**
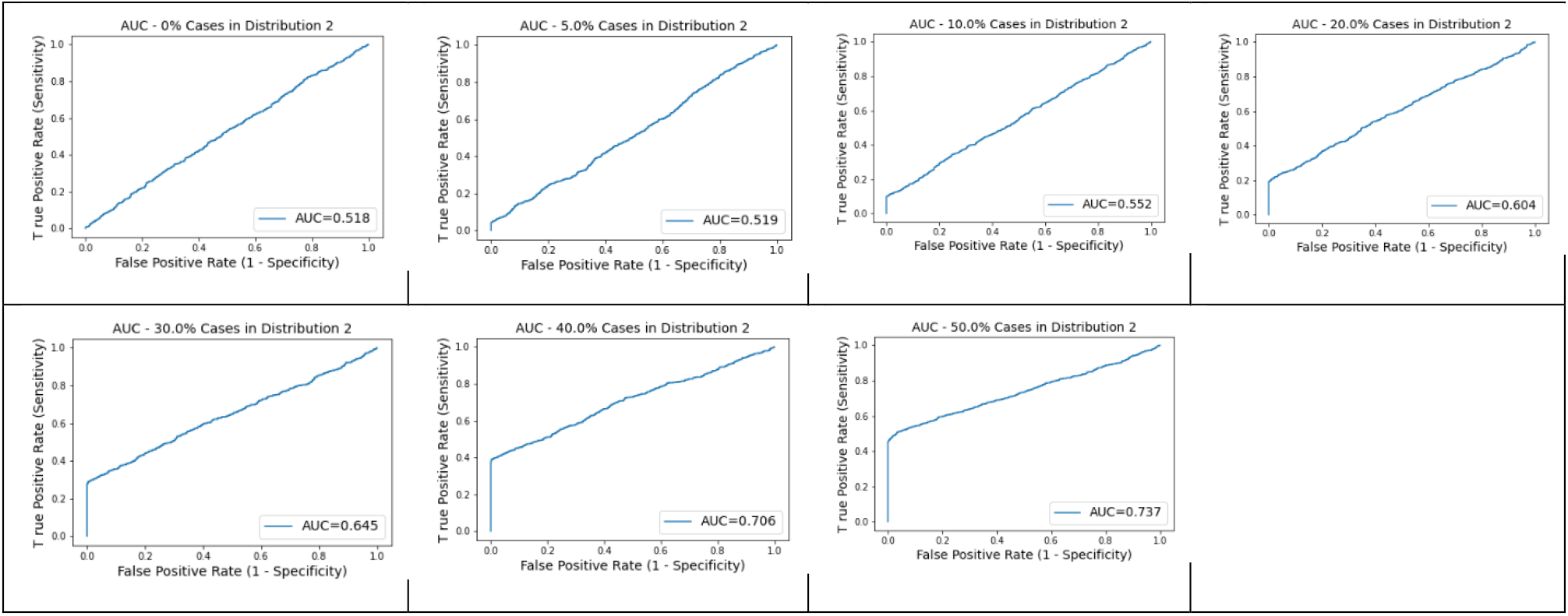
AUC plots for subtype groups of 0%, 5%, 10%, 20%, 30%, 40%, and 50% of the diseased cases. Random plots for each scenario are shown.

### Biological Data

We analyzed the AD gene expression data using FC, AUC, and BCD. Lists of the top six genes for each method are shown in **Table 2** and histograms for each of these genes are shown in **Figures 3, 4**, and **5**. Some of the FC and AUC plots exhibit tendency towards bimodality or increased skew, but in general they represent differences in expression across the majority of the samples, demonstrating their value for identifying biomarkers associated with large proportions of the cases. Across the 8650 genes, an AUC score of 0.74 represented a p-value ≤ 0.05.

**Table 2.** Top six genes for each analysis of the AD gene expression data.

| Fold Change |  | AUC |  | BCD |  |
| --- | --- | --- | --- | --- | --- |
| Gene | log2FC | Gene | AUC | Gene | BCD |
| GI_38201693-S | 1.448 | GI_4585642-S | 0.854 | GI_4502806-S | 0.403 |
| GI_40255112-S | 1.428 | GI_27734844-S | 0.830 | GI_17999536-S | 0.378 |
| GI_40254432-S | 1.342 | GI_24308166-S | 0.829 | GI_37540877-S | 0.378 |
| GI_40018630-A | 1.306 | GI_34577121-S | 0.822 | GI_28872783-A | 0.375 |
| GI_29744077-S | 1.288 | GI_13376557-S | 0.820 | GI_14589948-S | 0.374 |
| GI_27475984-S | 1.217 | GI_23312375-A | 0.815 | GI_23503234-A | 0.372 |

**Figure 3.**
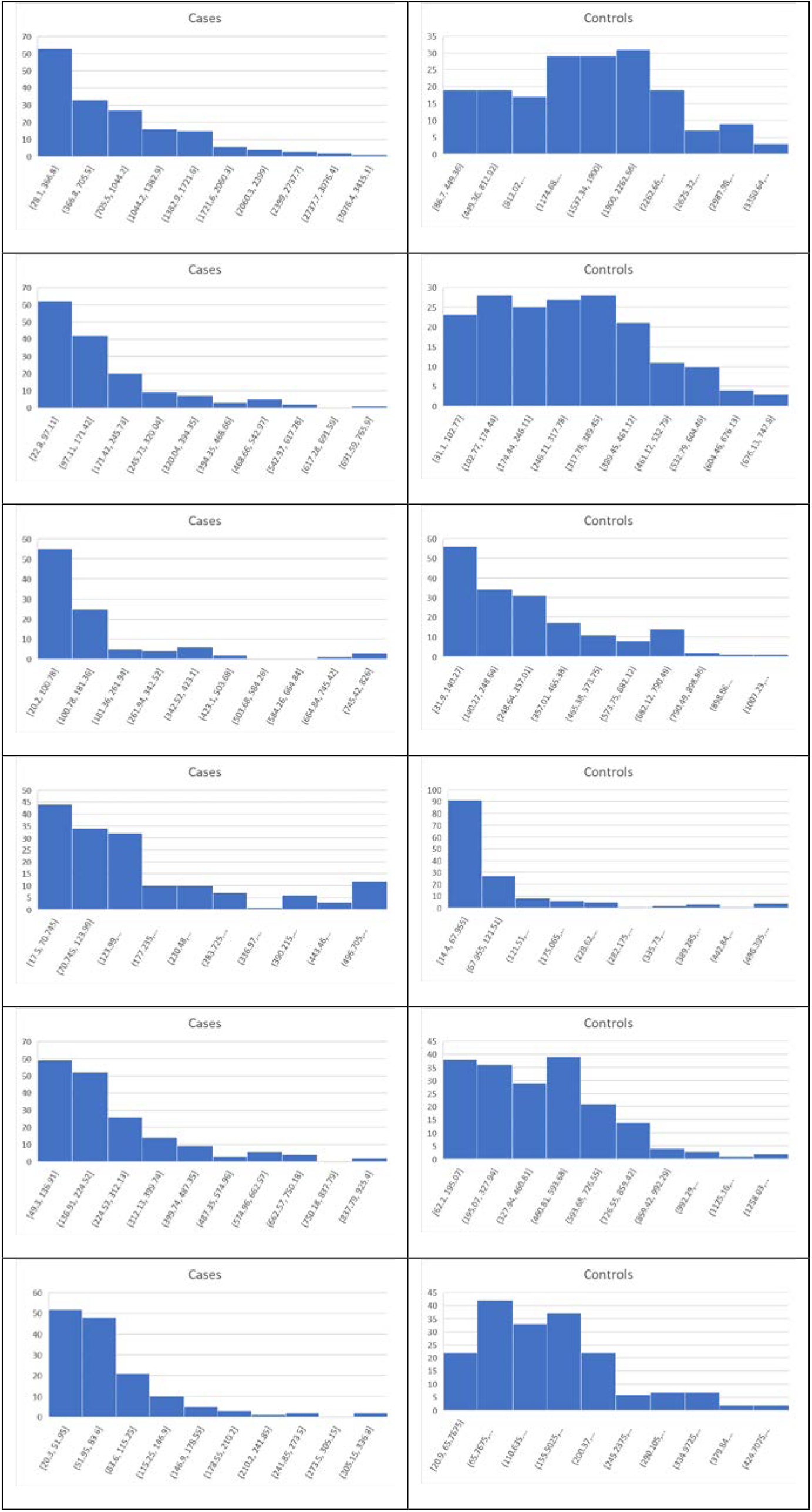
Histograms for the six top genes identified using FC. Each row corresponds to a gene in **Table 2** and are given in the same order.

**Figure 4.**
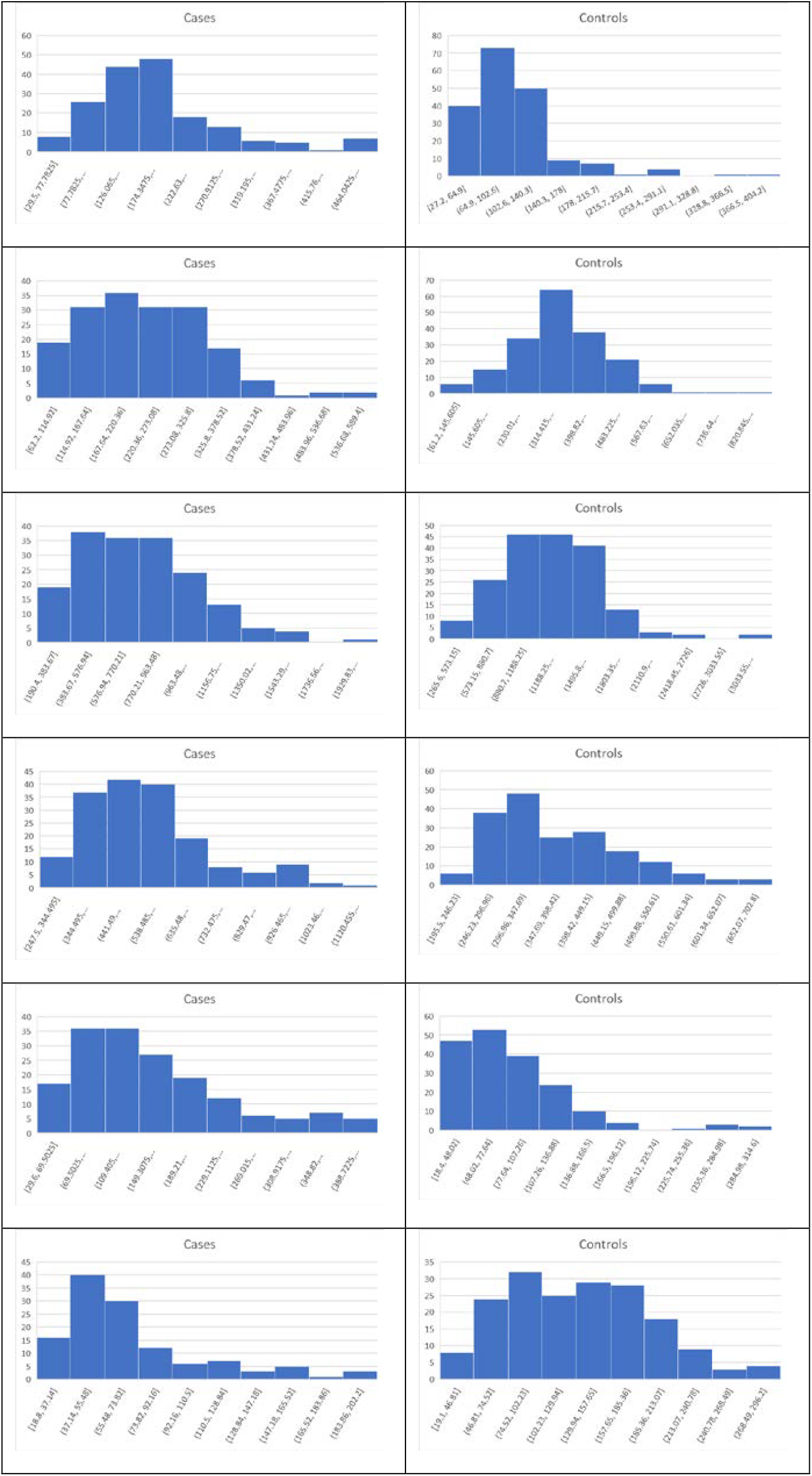
Histograms for the six top genes identified using AUC. Each row corresponds to a gene in **Table 2** and are given in the same order.

**Figure 5.**
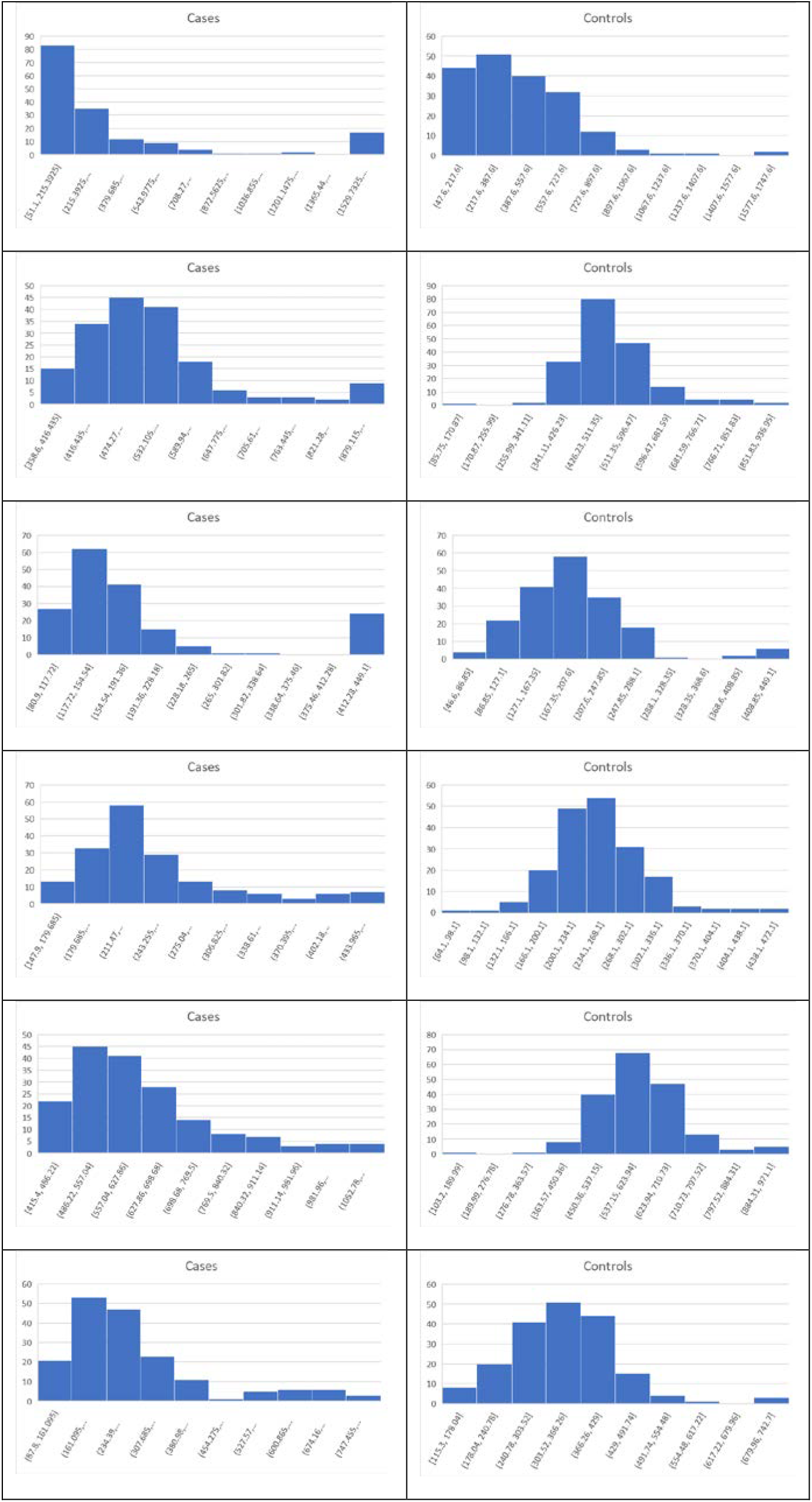
Histograms for the six top genes identified using BCD. Each row corresponds to a gene in **Table 2** and are given in the same order.

In our first round of BCD trials, the genes with the highest values proved to be spurious. For each of the highest BCD values, we extracted the covariate data for the samples in the second mode of the cases distribution. As shown in the **Supplementary Information**, more than x% of these second mode individuals were drawn from brain region 4, despite the fact that only 12.5% of the case samples overall were drawn from this region. Furthermore, only 4.8% of the control samples were drawn from region 4, yielding an imbalance of samples for this region. Consequently, diseased cases samples that were drawn from region 4 formed distinct subsets that created second modes for genes that were differentially expressed across the brain regions. These results demonstrate the power of BCD to identify subtypes, but do not yield information of interest regarding AD. As shown in the **Supplementary Information**, brain region 2 is also unbalanced between cases and controls.

In our second round of trials, we removed samples drawn from brain regions 2 and 4, yielding 137 AD cases and 175 normal controls. Histograms for the six genes with the highest BCD values are shown in **Figure 5**. These 8650 trials provided a cutoff value of 0.209 for p-value ≤ 0.05.

## Discussion

The simulation trials provided a comprehensive evaluation across methods with 1000 repetitions of large-scale trials comprised of 1000 cases and 1000 controls each and nearly ‘ideal’ subtype biomarkers representing each subtype percentage. The results from these trials are truly eye opening.

Assuming log2FC > 1 indicates significance, fold change performed extremely poorly. Even when 50% of the cases were associated with the subtype biomarker, the median log2FC value was only 0.608. The maximum across all 1000 trials was only 0.821. Consequently, all of the pseudo biomarkers would be thrown out, despite their nearly perfect discrimination.

It’s trickier to evaluate AUC, due to lack of a clear significance cutoff value. The literature points to 0.7 or 0.75 and our trials on AD gene expression provided a cutoff of 0.74 for p-value ≤ 0.05. All of the pseudo biomarkers had AUC values less than 0.7 across the 1000 trials for subsets less than 40%. Furthermore, the median for the 40% subset trial was 0.695. Significance emerged as the subset size grew to 50%.

Being a newly introduced metric, there are no established significance cutoffs for BCD. The AD gene expression data provided a cutoff value of 0.209 for p-value ≤ 0.05. Using this proxy, *every* one of the 1000 trials for subsets of 10% or more would be marked as significant.

As expected from the biological trials, the top six genes identified by FC and AUC show significant differences between the diseased cases and normal controls. While several of the top results exhibit some degree of bimodality, others tend towards differences across the majority of the samples. On the other hand, each top BCD result clearly delimitates a subgroup, without requiring differences for individuals that are not in the given subgroup.

A particularly interesting result is that the first round of BCD trials produced spurious associations for all of the top values due to the imbalance of cases and controls samples from brain region 4. While this imbalance, coupled with differential expression across brain regions, created subsets, none of these genes were included in the top six genes for FC or AUC.

It should be noted that BCD is not expected to identify global biomarkers. When all of the cases are associated with the biomarker, as has been shown for pTau-181 associations with AD, a shift in the cases mean, not modality, is expected. Shifts in means are directly captured by FC and ignored by BCD.

BCD enjoys the same favorable properties exhibited by AUC. No specific biomarker threshold or other parameters are utilized. The metric is simple and intuitive. Furthermore, examination of the corresponding histograms provides additional information beyond the scalar value and individuals representing the subtype are distinguished from those who are not associated.

At the same time, BCD does not suffer from AUC’s drawbacks. The lower bound on sample size is only limited by the ability to distinguish the bimodal coefficient for the distribution. Indeed, analytes that are already known to be unimodal under normal conditions do not necessarily require any new controls data.

AUC includes regions under the curve where analyte thresholds are not of practical interest and can be misleading when comparing two ROC curves that cross. Neither of these issues are of concern for BCD as the distributions of analyte levels, rather than TPRs and FPRs, dictate the computed values. Finally, high AUC value does not always correlate with a robust threshold for practical use of the biomarker. In contrast, high BCD value indicates strong bimodality, which corresponds to a natural inversion between the modes. While lower, but still significant, BCD values may indicate skewness rather than bimodality, inspection of the histogram will reveal the sensitivity of the threshold cutoff selected for diagnosis.

Precision medicine is based upon the assumption that different subtypes exist for the given disease. We show here that popular statistics used for assessing biomarkers, FC and AUC, generally perform suboptimally when heterogeneity exists. We also provide a new metric, BCD, which appears to hold promise in this domain.

## Data Availability

All data produced in the present study are available upon reasonable request to the authors

## Acknowledgements

Special thanks to Jamie Lea for insightful conversations and useful software. This research was funded by the Alzheimer’s Association (AARG-22-925002), National Institute on Aging (NIA) grants 1RF1AG053303-01 and 3RF1AG053303-01S2, and research grants from the University of Missouri – St. Louis.

